# Antibiotic Prophylaxis in Dental Implant Procedures: Evidence from a Systematic Review

**DOI:** 10.1101/2025.01.15.25320638

**Authors:** Qazi Jawad Hayat, Radaina Niaz, Naeem Ul Haq, Ziad Khan

**Affiliations:** Dentist, Postgraduate Resident, Oral and Maxillofacial Surgery - Khyber College of Dentistry, Peshawar, Pakistan; Dentist - Khyber College of Dentistry, Peshawar, Pakistan

**Keywords:** Implants, Dental implants, Antibiotic prophylaxis, Antibiotic resistance

## Abstract

**Background:** The use of antibiotic prophylaxis in dental implant surgery is controversial, with conflicting guidelines and variations in clinical practice. This systematic review aimed to evaluate the effectiveness of antibiotic prophylaxis in preventing implant failure and post-operative infection in healthy patients.

**Methods:** A systematic search of the PubMed database was conducted to identify relevant randomized controlled trials (RCTs). Four RCTs met the inclusion criteria and were included in the review. Data on implant failure, post-operative infection, and study characteristics were extracted.

**Results:** The four included RCTs, published between 2008 and 2022, encompassed 966 participants. The findings regarding the effect of antibiotic prophylaxis on implant failure were mixed. Two studies reported lower failure rates in the antibiotic group, one found similar rates, and one reported a higher failure rate in the antibiotic group. The findings regarding post-operative infections were also inconclusive, with generally low infection rates reported across both groups.

**Conclusion:** The evidence regarding the benefit of antibiotic prophylaxis in preventing implant failure and post-operative infection in healthy patients is mixed and inconclusive. Further well-designed and adequately powered RCTs are needed to clarify the role of antibiotic prophylaxis in this setting. In the meantime, clinicians should carefully consider the individual patient’s risk factors, the potential benefits and harms of antibiotic prophylaxis, and the available evidence when making decisions about antibiotic use in dental implant surgery.

## Introduction

Antibiotics are antimicrobial agents effective against bacteria, acting to inhibit bacterial growth or induce bacterial death, thereby facilitating elimination by the host’s immune system ^1.^ However, the widespread and often inappropriate use of antibiotics has contributed to the emergence and spread of antibiotic-resistant bacteria, posing a significant global health threat ^2^. Prudent antibiotic use is therefore essential to mitigate this growing problem.

Dental implant placement has become a standard treatment modality for replacing missing teeth, whether due to extraction or congenital absence. Its popularity stems from high reported success rates and long-term stability ^3^. Preoperative antibiotic prophylaxis is frequently prescribed before implant placement with the aim of preventing early implant failure. However, current guidelines regarding this practice are inconsistent. Notably, discrepancies exist between recommendations from UK professional bodies such as the Faculty of General Dental Practice (FGDP) and the Faculty of Dental Surgery of the Royal College of Surgeons of England (FDS), with some guidelines advising against routine antibiotic use for implant placement ^4^.

Conflicting evidence further complicates this issue. While some studies, such as that by Anitua et al. (2009), suggest that antibiotic prophylaxis may not be necessary for single implant placement, others, such as Esposito et al. (2013), have proposed it as essential ^5–6^. This lack of consensus underscores the need for a comprehensive evaluation of the existing evidence.

Therefore, this systematic review aims to evaluate and synthesize the available evidence on the effectiveness and necessity of antibiotic prophylaxis in preventing post-operative complications, including implant failure and promoting overall implant success, in healthy patients undergoing dental implant procedures.

## Material and Methods

This systematic review was conducted in accordance with the Preferred Reporting Items for Systematic Reviews and Meta-Analyses (PRISMA) 2020 statement ^7^. The primary research question guiding this review was: Given the rising concern over antibiotic resistance, how does the use of antibiotic prophylaxis affect the prevention of post-operative complications in healthy patients undergoing dental implant procedures compared to no antibiotic prophylaxis? This was further specified using the PICO framework:

Population (P): Healthy patients undergoing dental implant procedures (without comorbidities known to significantly increase infection risk or impair (osseointegration).

Intervention (I): Preoperative administration of antibiotic prophylaxis.

Comparison (C): No antibiotic prophylaxis (placebo or no intervention).

Outcomes (O): Implant failure, implant success, and post-operative infection.

Search Strategy:

A systematic literature search conducted in the PubMed (MEDLINE) database to identify relevant studies published up to January 2025. The search strategy utilized a combination of keywords and Medical Subject Headings (MeSH terms), including: “dental implant,” “implants,” “prophylaxis,” “prophylactic antibiotics,” and “antibiotics.” Various combinations of these terms, using Boolean operators (AND, OR, NOT), employed to maximize search sensitivity.

### Study Selection

Two independent reviewers screened titles and abstracts of retrieved records based on pre-defined inclusion and exclusion criteria. Full texts of potentially eligible studies then retrieved and independently assessed by the same two reviewers. Discrepancies at both stages were resolved through discussion or consultation with a third reviewer. The study selection process was documented using a PRISMA flow diagram.

### Inclusion Criteria

Participants: Systemically healthy adult patients undergoing dental implant placement. Systemic health will be defined as the absence of comorbidities known to significantly increase the risk of infection or impair osseointegration (e.g., uncontrolled diabetes mellitus, immunocompromising conditions, active infections, history of bisphosphonate use, head/neck radiation).

Intervention: Preoperative administration of antibiotic prophylaxis (any type, dosage, route, timing within 1 hour of surgery) given explicitly for prophylaxis.

Comparison: Control group receiving placebo or no intervention. Study Design: Randomized controlled trials (RCTs).

Publication Characteristics: English language, available full text.

### Exclusion Criteria

Patients with systemic conditions increasing infection risk/impairing osseointegration. Concomitant interventions confounding prophylaxis assessment.

Studies primarily comparing different antibiotic types/dosages/routes.

Non-RCT study designs.

Non-English language or unavailable full text.

### Data Extraction

A standardized data extraction form was developed. Two independent reviewers extracted data from included studies (n = 4), with discrepancies resolved by discussion. The following data were extracted, corresponding to the fields in the data extraction table 1. Data were recorded in a pre-designed spreadsheet structured to facilitate data analysis. Any discrepancies encountered during data extraction were documented and resolved through discussion.

**Table 1:**
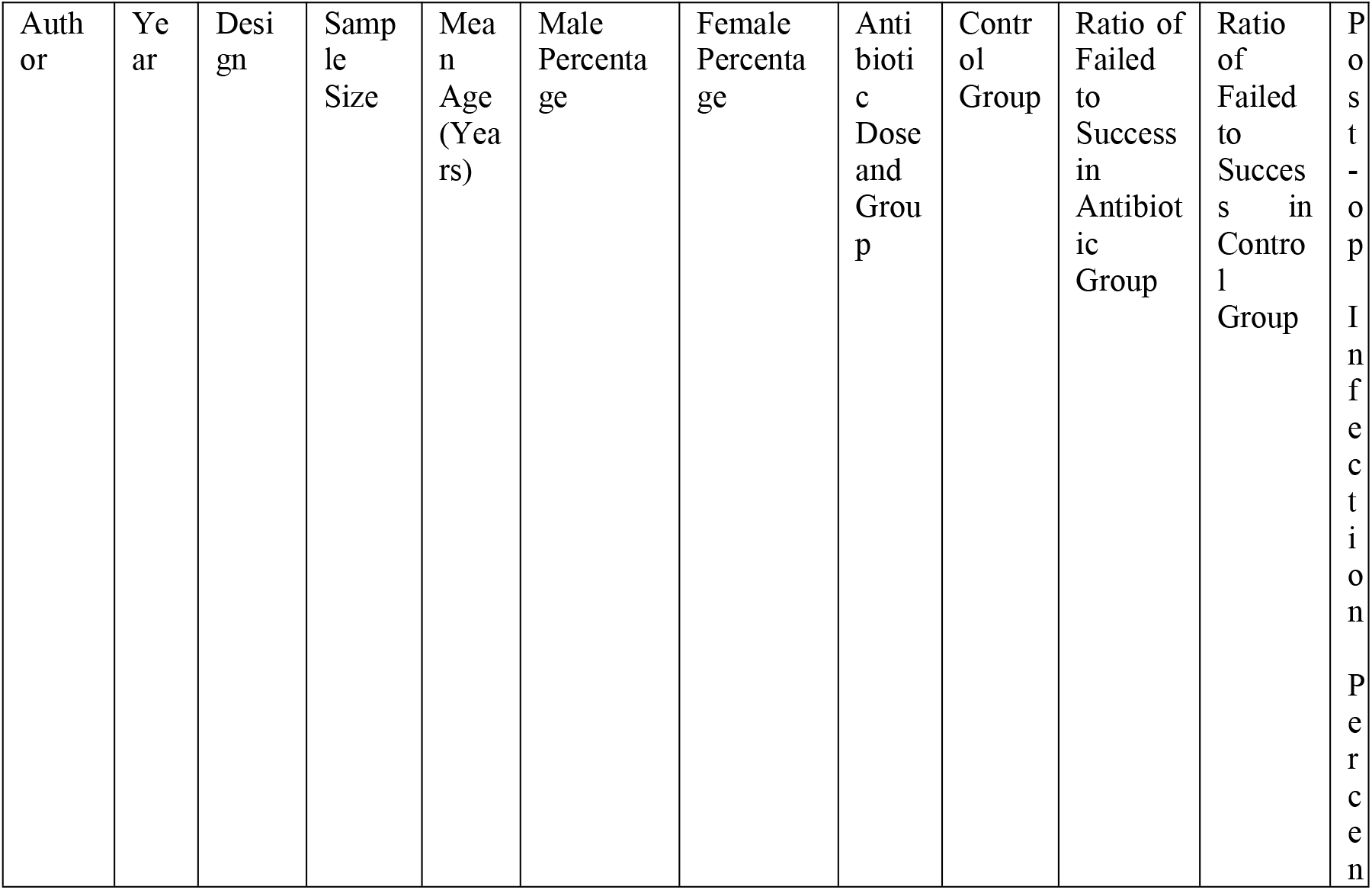
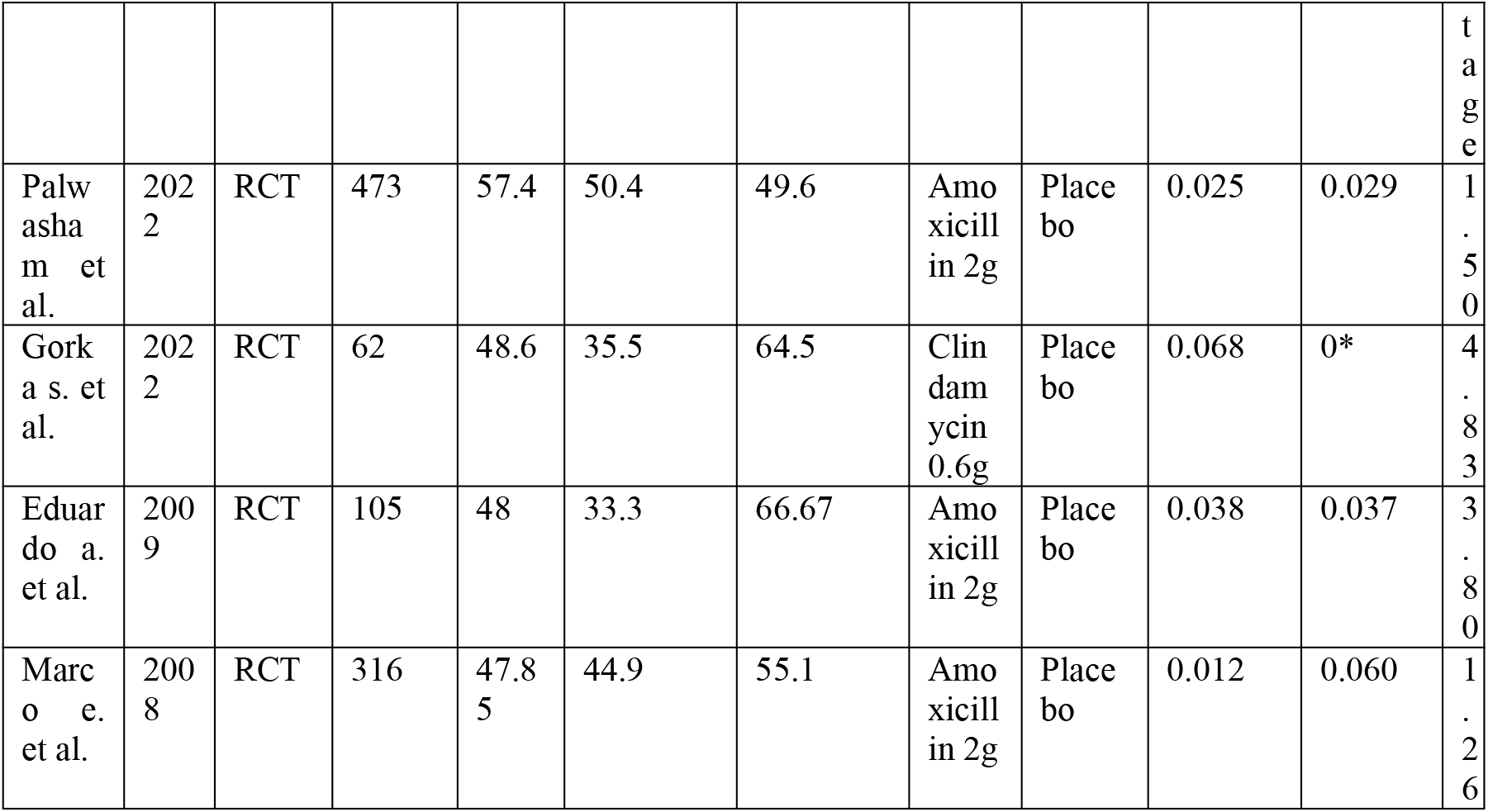
Characteristics of Included Studies.

## Results

### 1. Study Selection

The electronic search of PubMed yielded a large number of records. After removing duplicates and screening titles and abstracts, articles were assessed for full-text eligibility. After application of inclusion and exclusion criteria, four randomized controlled trials (RCTs) met the inclusion criteria and were included in this systematic review (Figure 1).

**Figure 10:**
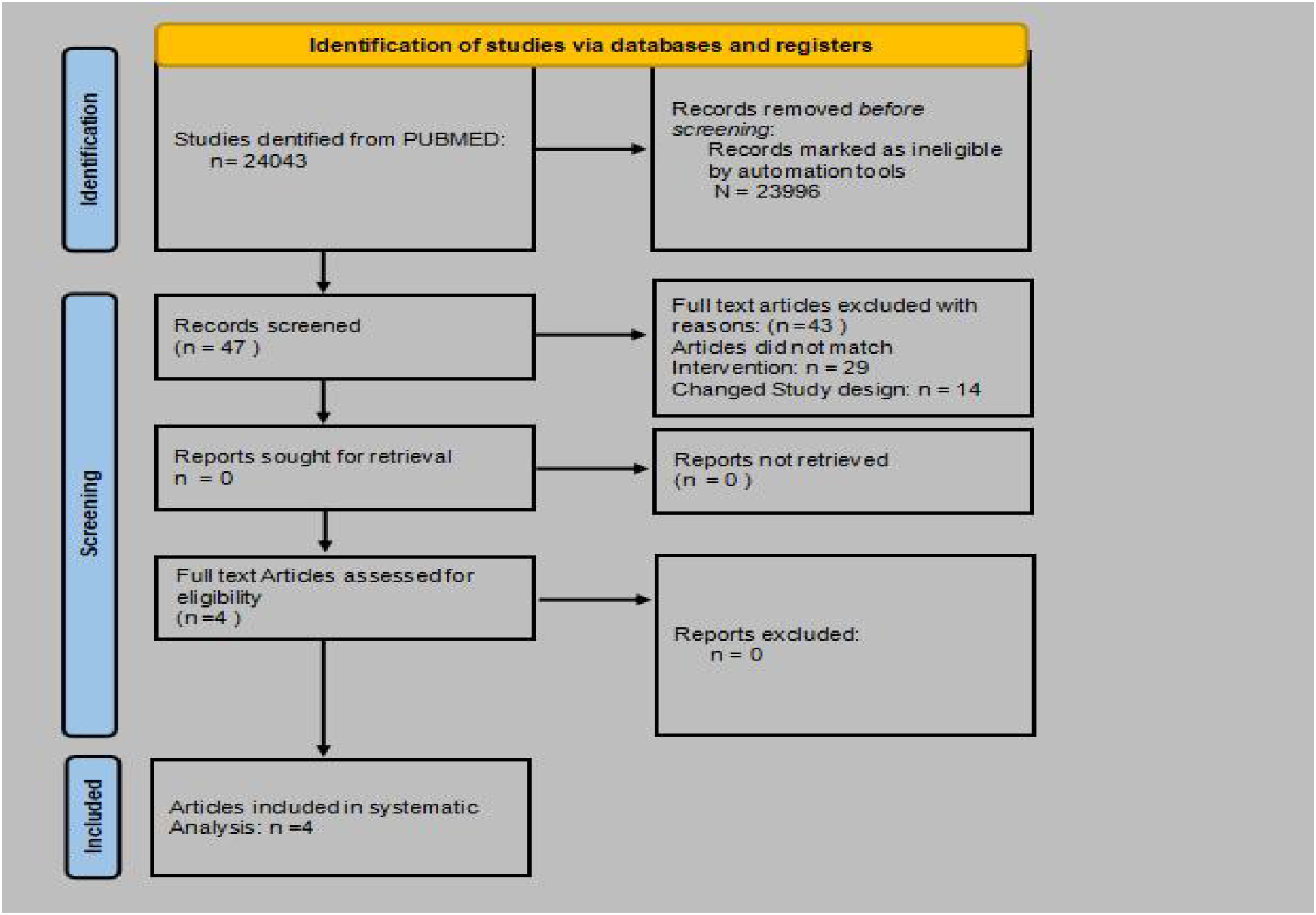
PRISMA Flow Chart for Studies.

### 2. Study Characteristics

The characteristics of the four included studies are summarized in Table 1. All studies were RCTs published between 2008 and 2022. The sample sizes ranged from 62 to 473 participants. The mean ages of participants ranged from 47.85 to 57.4 years. All studies compared preoperative antibiotic prophylaxis with a placebo control.

### 3. Risk of Bias Assessment

The risk of bias assessment using the Cochrane Risk of Bias 2 tool suggested a generally low risk of bias across the included studies. There were some minor concerns regarding blinding in some studies, as full details of blinding procedures were not always reported.

### 4. Synthesis of Results

This section is organized by outcome: implant failure and post-operative infection.

#### 4.1. Implant Failure

Four studies reported on implant failure. Palwasha m et al. (2022) reported a low failure rate in both the antibiotic (2.5%) and placebo (2.9%) groups. Gorka s. et al. (2022) observed a higher failure rate in the clindamycin group (6.8%) compared to no failures in the placebo group. Eduardo a. et al. (2009) reported similar failure rates in the antibiotic (3.8%) and placebo (3.7%) groups. Marco e. et al. (2008) observed a lower failure rate in the antibiotic group (1.2%) compared to the placebo group (6%). Overall, the findings regarding the effect of antibiotic prophylaxis on implant failure are mixed. While some studies suggest a potential benefit of antibiotics, particularly the study by Marco e. et al. (2008), others show no clear difference or even a trend towards higher failure rates in the antibiotic group (Gorka s. et al., 2022).

#### 4.2. Post-operative Infection

All four studies reported on post-operative infection rates. The reported infection rates were generally low across both groups. Palwasha m et al. (2022) reported a 1.5% infection rate, Gorka s. et al. (2022) reported 4.83%, Eduardo a. et al. (2009) reported 3.8%, and Marco e. et al. (2008) reported 1.26%. Due to variations in reporting and the small number of events, it is difficult to draw firm conclusions about the effect of antibiotic prophylaxis on post-operative infection.

## Discussion

This systematic review aimed to evaluate the effectiveness and necessity of antibiotic prophylaxis in preventing post-operative complications, specifically implant failure and infection, in healthy patients undergoing dental implant procedures. By synthesizing the available evidence from randomized controlled trials (RCTs), this review sought to provide clarity on a contentious issue in dental practice, where guidelines and clinical practice vary.

## Summary of Findings

The review included four RCTs published between 2008 and 2022, encompassing a total of 966 participants. The studies varied in sample size, ranging from 62 to 473 participants, with mean ages ranging from 47.85 to 57.4 years. All studies compared preoperative antibiotic prophylaxis with a placebo control, with amoxicillin being the most frequently used antibiotic.

The findings regarding the effect of antibiotic prophylaxis on implant failure were mixed. Two studies reported lower failure rates in the antibiotic group compared to the placebo group, one study found similar failure rates between the two groups, and one study reported a higher failure rate in the antibiotic group. The findings regarding the effect of antibiotic prophylaxis on post-operative infections were also inconclusive, with generally low infection rates reported across both antibiotic and placebo groups.

## Strengths and Limitations

This systematic review has several strengths. It adhered to a rigorous methodology, including a comprehensive search strategy, strict inclusion and exclusion criteria, and a systematic assessment of the risk of bias of included studies. The review also included only RCTs, which are considered the gold standard for evaluating the effectiveness of interventions.

However, the review also has some limitations. The small number of included studies limits the statistical power of the review and the generalizability of the findings. The heterogeneity in study design, patient populations, and outcome reporting also limits the ability to draw firm conclusions. Additionally, the risk of bias assessment revealed some concerns regarding blinding in some of the included studies, which may have influenced the results.

## Implications for Clinical Practice

The findings of this review have implications for clinical practice. Given the mixed and inconclusive evidence regarding the benefit of antibiotic prophylaxis in preventing implant failure and post-operative infections in healthy patients, the routine use of antibiotic prophylaxis for dental implant placement cannot be definitively recommended or discouraged.

Clinicians should carefully consider the individual patient’s risk factors, the potential benefits and harms of antibiotic prophylaxis, and the available evidence when making decisions about antibiotic use in dental implant surgery. Shared decision-making with patients, incorporating their preferences and values, is also essential.

## Implications for Future Research

This systematic review highlights the need for further well-designed and adequately powered RCTs to clarify the role of antibiotic prophylaxis in dental implant surgery. Future studies should address the limitations of the current evidence by:

- Including larger sample sizes to increase statistical power
- Using standardized outcome measures and reporting to reduce heterogeneity
- Implementing rigorous blinding procedures to minimize bias
- Evaluating the cost-effectiveness of antibiotic prophylaxis
- Assessing the long-term effects of antibiotic prophylaxis on implant outcomes and the development of antibiotic resistance

## Conclusion

This systematic review found mixed and inconclusive evidence regarding the effectiveness of antibiotic prophylaxis in preventing implant failure and post-operative infections in healthy patients undergoing dental implant procedures. The small number of included studies, heterogeneity in study design and outcome reporting, and concerns about bias limit the strength of the conclusions. Further well-designed and adequately powered RCTs are needed to clarify the role of antibiotic prophylaxis in this setting.

## Data Availability

All data produced in the present work are contained in the manuscript

